# Projections and early-warning signals of a second wave of the COVID-19 epidemic in Illinois

**DOI:** 10.1101/2020.07.06.20147868

**Authors:** Zachary J. Weiner, George N. Wong, Ahmed Elbanna, Alexei V. Tkachenko, Sergei Maslov, Nigel Goldenfeld

## Abstract

We present two different scenarios for a second wave of the COVID-19 epidemic in Illinois and simulate them using our previously described age-of-infection model, calibrated to real-time hospital and deaths data. In the first scenario we assume that the parameters of the second wave in Illinois would be similar to those currently observed in other states such as Arizona, Florida, and Texas. We estimate doubling times of hospitalizations and test positivity in all states with relevant publicly available data and calculate the corresponding effective reproduction numbers for Illinois. These parameters are remarkably consistent in states with rapidly growing epidemics. We conjecture that the emergence of the second wave of the epidemic in these states can be attributed to superspreading events at large parties, crowded bars, and indoor dining. In our second, more optimistic scenario we assume changes in Illinois state policy would result in successful mitigation of superspreading events and thus would lower the effective reproduction number to the value observed in late June 2020. In this case our calculations show effective suppression of the second wave in Illinois. Our analysis also suggests that the logarithmic time derivatives of COVID-19 hospitalizations and case positivity can serve as a simple but strong early-warning signal of the onset of a second wave.

---

A number of regions within the United States are currently experiencing second waves following their reopenings. We consider whether or not the transition to Restore Illinois Phase 4^1^ will lead to a second wave, and if so, how bad will it be. While there is anecdotal evidence that the second waves are in part caused by people failing to social distance in bars, at parties, etc., it is not easy to relate these social distancing failures to parameters in complicated epidemiological models. It is thus challenging to construct a reliable and actionable prediction for the state of Illinois.

To circumvent this difficulty, we use fits to the current exponential growth rates of epidemic dynamics in affected states to estimate the corresponding Phase 4 scenarios for Illinois. In lieu of performing computationally expensive calibration of detailed epidemiological models for these states (as we perform for Illinois), we carried out an approximate calculation of the doubling time of hospitalizations (i.e., the current number of hospital beds occupied by COVID-19 patients) and test positivity rates in Texas and Arizona and of the test positivity rate in Florida. The resulting estimates for Florida, Arizona, and Texas are remarkably consistent, with the percentage by which *R*_*t*_ relaxes towards *R*_0_ ranging between 22% and 31%. We then model scenarios for Phase 4 in Illinois designed to reproduced the currently-observed growth rates in these states (which have already relaxed social distancing strategies in line with those introduced by Illinois’s Phase 4), using our age-of-infection model of COVID-19 dynamics in Illinois [2]. We contrast the second wave exhibited by these results with an additional scenario in which Illinois subsequently reverts the policy changes enacted by Phase 4.

As a preamble to our estimate, we point out that there is a suitable “early-warning signal” that the state of Illinois and other states could use to monitor the second wave: the rate of change of case positivity, which is arguably a leading indicator compared to hospitalization, ICU, and death data. The data in Figure 1 shows positivity rates for Arizona, Florida and Texas, with Illinois shown for comparison. Currently, the Illinois Department of Public Health (IDPH) sets a case positivity of 10% as a threshold for a warning level on a per-county basis.^2^ This is not a good metric to use because it depends on an arbitrary value for the base level of case positivity. Arizona had a baseline level of about 8% before the second wave started, and so it quickly passed the 10% value. Florida and Texas, however, started at lower baseline levels and so the momentum of the second wave was clearly visible a week or so before they breached the 10% threshold.

**FIG. 1.**
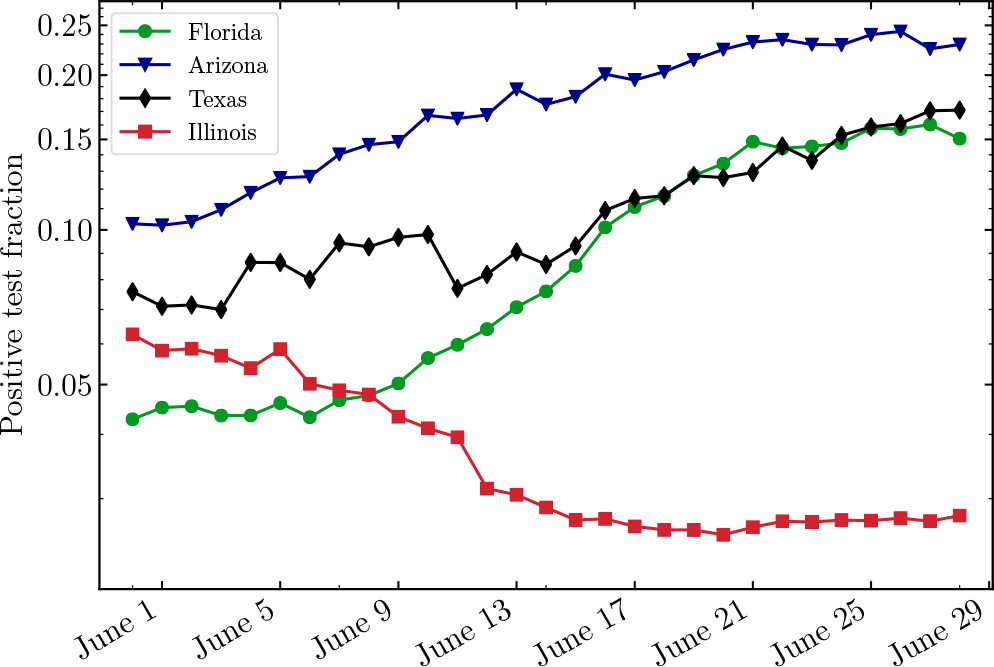
Daily COVID-19 positivity rate in selected US states: Illinois (red squares), Texas (black diamonds), Arizona (blue triangles), and Florida (green circles). To reduce noise, the data are smoothed by a 7-day moving average.

Thus, action could have been taken earlier to prevent the situations in these states from getting out of control.

## The second wave across the US quantified by growth rates in hospitalizations and test positivity

In Illinois, the early epidemic exhibited a basic reproduction number *R*_0_ = 2.32 as estimated using our epidemiological model [2]. This corresponds to an exponential growth rate g(early in IL) = 0.272, i.e., a doubling time of ln(2)/0.272 = 2.5 days.

Our age-of-infection model [2] assumes a Gamma-distributed serial interval with a mean *τ*_*s*_ = 4 days and a standard deviation *σ*_*s*_ = 3.25 days. In this case the effective reproduction number *R*_*t*_ is related to the exponential growth (or decay) rate *g* via

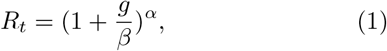

where 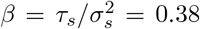 and 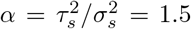 are the rate and the shape parameters of the Gamma distribution of serial intervals (see Eq. 2.9 of Ref. [4] for a derivation).

In our model *R*_*t*_ = *R*_0_*· M* (*t*) *· S*(*t*), where *M* (*t*) is a mitigation multiplier accounting for the effects of non-pharmaceutical intervention like social distancing, and *S*(*t*) is the fraction of susceptible population at time *t*. We assume that the transition to Phase 4 results in partial relaxation of *M* (*t*) toward unity (which would correspond to a completely unmitigated epidemic). Our simulations predict that by the time of transition to Phase 4 the susceptible fraction of Illinois’s population was around *S*(Phase 3) = 0.91. That is, by the beginning of Phase 4 roughly 9% of the population have been infected by COVID-19; in our model we assume previously infected individuals remain immune for the duration of the simulation. We model the transition from Phase 3 to Phase 4 by increasing the mitigation factor from *R*_*t*_(Phase 3) = 0.92 to a higher value *R*_*t*_(Phase 4). On the other hand, a complete relaxation of the mitigation factor to 1 would increase *R*_*t*_ from *R*_*t*_(Phase 3) = 0.92 all the way up to *R*_0_ · *S*(Phase 3). The extent of the relaxation of mitigation is quantified by the parameter *E*:

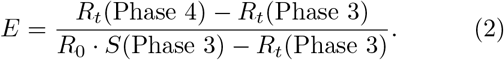

### Texas

Figure 2 shows COVID-19 hospitalizations (purple) and the daily case positivity rate (green) in Texas between June 1 and June 28 as reported by the COVID Tracking Project [5]. Over the period from June 15 through June 28 hospitalizations in Texas have increased with an exponential growth rate *g*(TX) = 0.068 corresponding to a ln(2)*/*0.068 = 10.2 day doubling time.

**FIG. 2.**
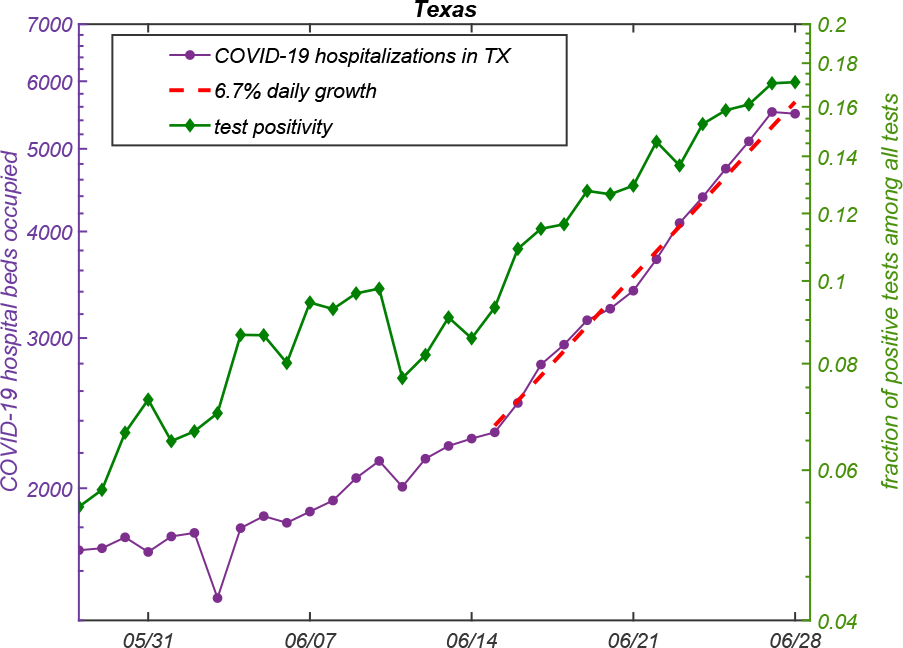
The exponential growth in COVID-19 hospitalizations (purple, left side) and daily case positivity rate (green, right side) in Texas. Hospitalizations between June 15 and June 28 have daily growth rate of around 6.8%. To reduce noise 7-day moving average has been applied to the test positivity rate. The ranges of the left and the right sides of the plot were selected in such a way that equal exponential growth rates of hospitalizations and test positivity would correspond to equal slopes.

Applying Eq. 1 with *g*(TX) = 0.068 observed in the second wave in Texas, one obtains *R*_*t*_(Phase 4 in IL matching TX) = 1.28.

For *R*_*t*_(Phase 4 matching TX) = 1.28, Eq. 2 gives the relaxation factor *E*(Phase 4 matching TX) = 31%.

### Arizona

Arizona shows somewhat slower exponential growth in hospitalizations between June 15 and June 28, 2020 (see Figure 3). The daily growth rate is *g*(AZ) = 0.045 (4.5%) corresponding to a doubling time of ln(2)*/*0.45 ≈ 15.4 days. In this case, Eq. 1 gives *R*_*t*_(Phase 4 matching AZ) = 1.19 corresponding to the *E*(Phase 4 matching AZ) = 22% (see Eq. 2).

**FIG. 3.**
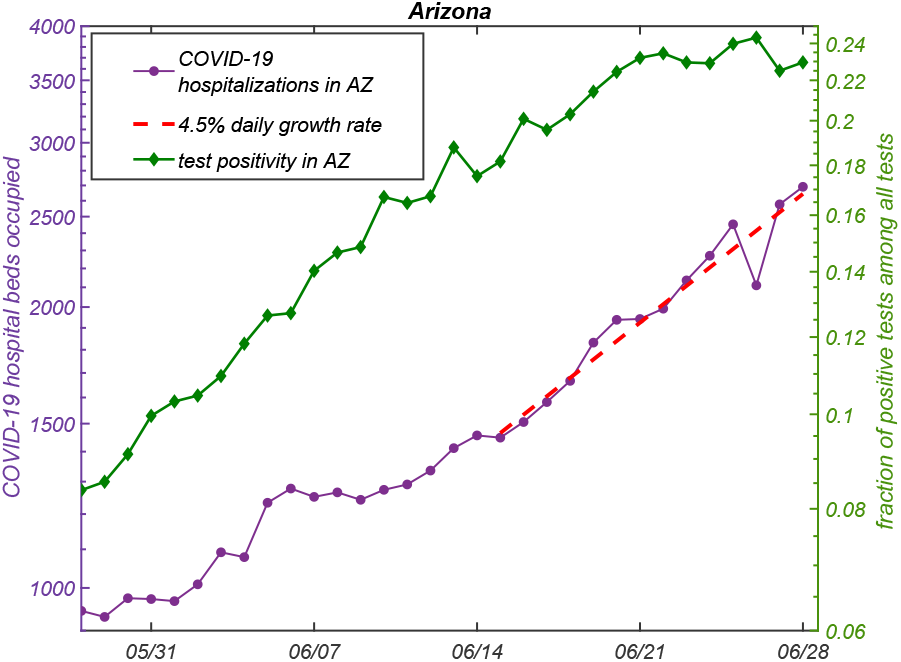
The exponential growth in COVID-19 hospitalizations (purple, left side) and daily case positivity rate (green, right side) in Arizona. Hospitalizations between June 15 and June 28 have daily growth rate of around 4.5% (red dashed line). To reduce noise 7-day moving average has been applied to the test positivity rate. The ranges of the left and the right sides of the plot were selected in such a way that equal exponential growth rates of hospitalizations and test positivity would correspond to equal slopes.

**FIG. 4.**
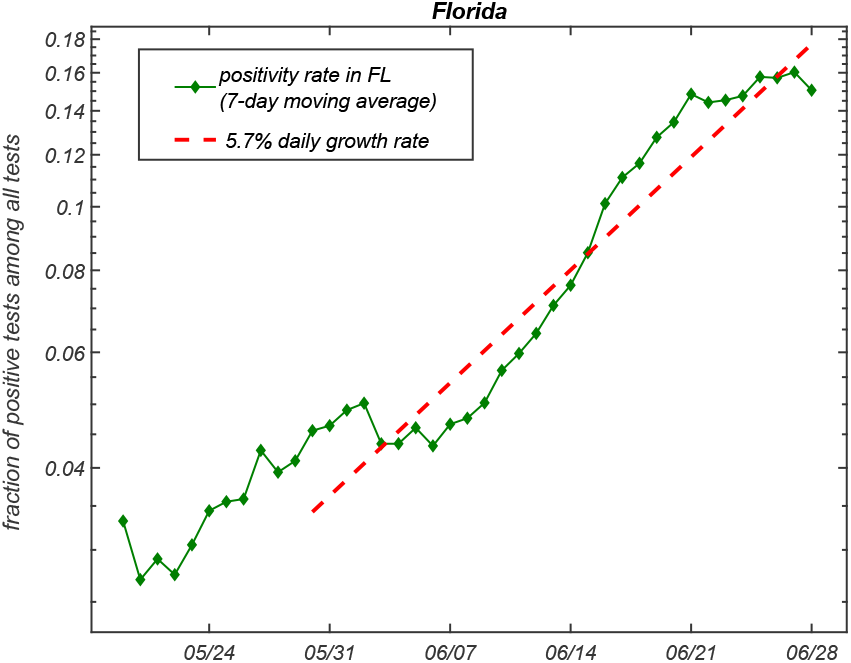
The exponential growth in COVID-19 test positivity rate (green) in Florida has average daily growth rate about 5.7% between May 30 and June 28 (red dashed line). To reduce noise 7-day moving average has been applied to the test positivity rate.

The growth rate of the test positivity in Arizona is similar to that of hospitalizations throughout most of the time window shown in Figure 3 except at the very end of the interval shown in Figure 3, where it starts to flatten out.

### Florida

The COVID Tracking Project [5] does not have hospitalization data for Florida. To obtain an alternative estimate of recent exponential growth in Florida, we instead use its reported test positivity rate, which we find grows exponentially with daily growth rate *g*(FL) = 0.057 (5.7% or a doubling time of 12.2 days). between May 30 and June 28. Translating these estimates to Phase 4 in Illinois via Eq. 1 gives *R*_*t*_(Phase 4 matching FL) = 1.24 and *E*(Phase 4 matching FL) = 27%.

### Other states

In Table I, we summarize the exponential growth (or decay) rates of hospitalizations (first column) and test positivity (last column) for all states for which the COVID Tracking Project [5] has relevant information. We report the slope of the linear regression to the logarithm of the corresponding variables (hospitalizations and the 7-day moving average of test positivity) over a two week period ending on June 28. The states are ranked in the order of decreasing hospitalization growth rates.

**TABLE I.**
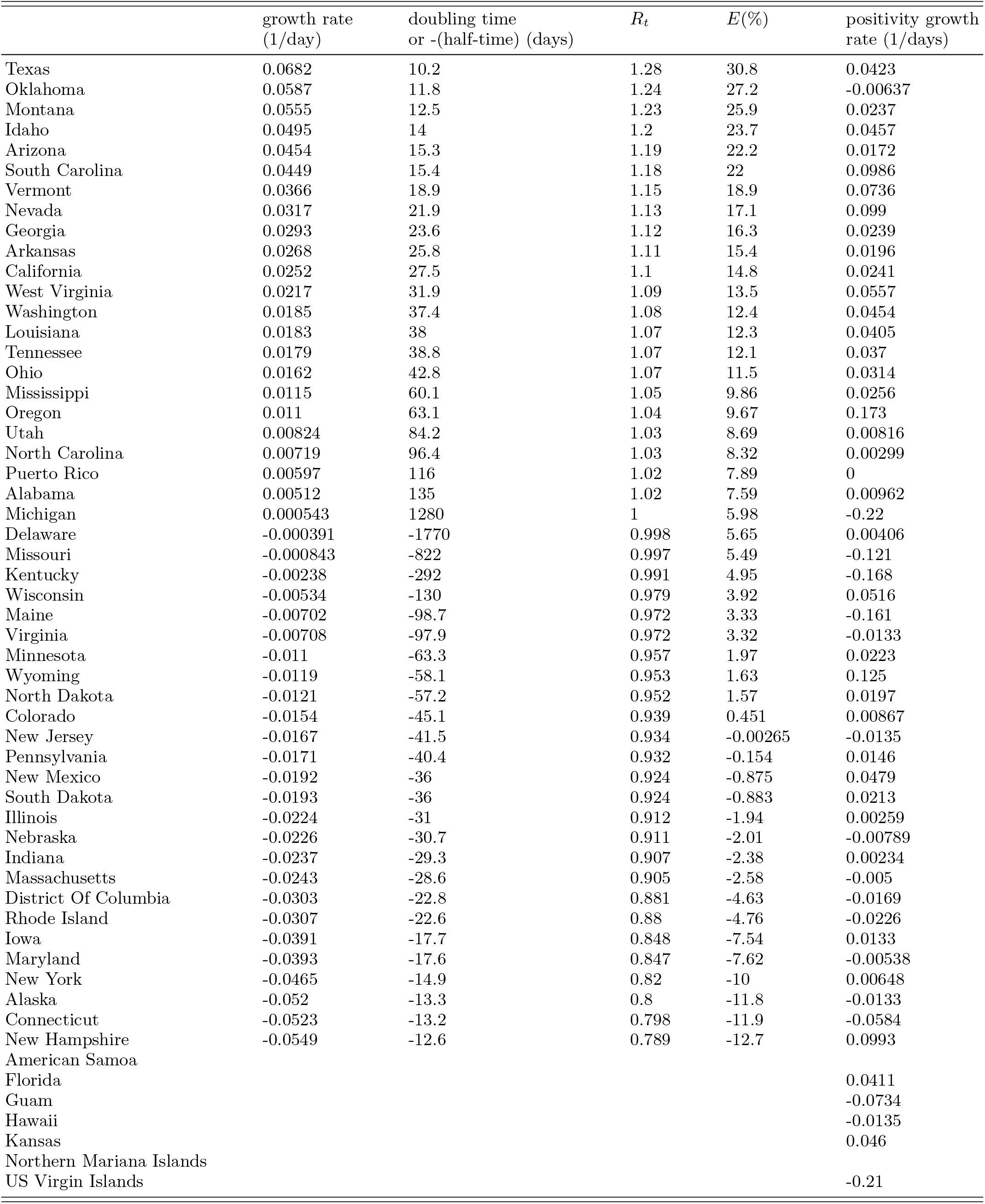

| | growth rate<br>(1/day) | doubling time<br>or -(half-time) (days) | $R_t$ | $E(\%)$ | positivity growth<br>rate (1/days) |
| --- | --- | --- | --- | --- | --- |
| Texas | 0.0682 | 10.2 | 1.28 | 30.8 | 0.0423 |
| Oklahoma | 0.0587 | 11.8 | 1.24 | 27.2 | -0.00637 |
| Montana | 0.0555 | 12.5 | 1.23 | 25.9 | 0.0237 |
| Idaho | 0.0495 | 14 | 1.2 | 23.7 | 0.0457 |
| Arizona | 0.0454 | 15.3 | 1.19 | 22.2 | 0.0172 |
| South Carolina | 0.0449 | 15.4 | 1.18 | 22 | 0.0986 |
| Vermont | 0.0366 | 18.9 | 1.15 | 18.9 | 0.0736 |
| Nevada | 0.0317 | 21.9 | 1.13 | 17.1 | 0.099 |
| Georgia | 0.0293 | 23.6 | 1.12 | 16.3 | 0.0239 |
| Arkansas | 0.0268 | 25.8 | 1.11 | 15.4 | 0.0196 |
| California | 0.0252 | 27.5 | 1.1 | 14.8 | 0.0241 |
| West Virginia | 0.0217 | 31.9 | 1.09 | 13.5 | 0.0557 |
| Washington | 0.0185 | 37.4 | 1.08 | 12.4 | 0.0454 |
| Louisiana | 0.0183 | 38 | 1.07 | 12.3 | 0.0405 |
| Tennessee | 0.0179 | 38.8 | 1.07 | 12.1 | 0.037 |
| Ohio | 0.0162 | 42.8 | 1.07 | 11.5 | 0.0314 |
| Mississippi | 0.0115 | 60.1 | 1.05 | 9.86 | 0.0256 |
| Oregon | 0.011 | 63.1 | 1.04 | 9.67 | 0.173 |
| Utah | 0.00824 | 84.2 | 1.03 | 8.69 | 0.00816 |
| North Carolina | 0.00719 | 96.4 | 1.03 | 8.32 | 0.00299 |
| Puerto Rico | 0.00597 | 116 | 1.02 | 7.89 | 0 |
| Alabama | 0.00512 | 135 | 1.02 | 7.59 | 0.00962 |
| Michigan | 0.000543 | 1280 | 1 | 5.98 | -0.22 |
| Delaware | -0.000391 | -1770 | 0.998 | 5.65 | 0.00406 |
| Missouri | -0.000843 | -822 | 0.997 | 5.49 | -0.121 |
| Kentucky | -0.00238 | -292 | 0.991 | 4.95 | -0.168 |
| Wisconsin | -0.00534 | -130 | 0.979 | 3.92 | 0.0516 |
| Maine | -0.00702 | -98.7 | 0.972 | 3.33 | -0.161 |
| Virginia | -0.00708 | -97.9 | 0.972 | 3.32 | -0.0133 |
| Minnesota | -0.011 | -63.3 | 0.957 | 1.97 | 0.0223 |
| Wyoming | -0.0119 | -58.1 | 0.953 | 1.63 | 0.125 |
| North Dakota | -0.0121 | -57.2 | 0.952 | 1.57 | 0.0197 |
| Colorado | -0.0154 | -45.1 | 0.939 | 0.451 | 0.00867 |
| New Jersey | -0.0167 | -41.5 | 0.934 | -0.00265 | -0.0135 |
| Pennsylvania | -0.0171 | -40.4 | 0.932 | -0.154 | 0.0146 |
| New Mexico | -0.0192 | -36 | 0.924 | -0.875 | 0.0479 |
| South Dakota | -0.0193 | -36 | 0.924 | -0.883 | 0.0213 |
| Illinois | -0.0224 | -31 | 0.912 | -1.94 | 0.00259 |
| Nebraska | -0.0226 | -30.7 | 0.911 | -2.01 | -0.00789 |
| Indiana | -0.0237 | -29.3 | 0.907 | -2.38 | 0.00234 |
| Massachusetts | -0.0243 | -28.6 | 0.905 | -2.58 | -0.005 |
| District Of Columbia | -0.0303 | -22.8 | 0.881 | -4.63 | -0.0169 |
| Rhode Island | -0.0307 | -22.6 | 0.88 | -4.76 | -0.0226 |
| Iowa | -0.0391 | -17.7 | 0.848 | -7.54 | 0.0133 |
| Maryland | -0.0393 | -17.6 | 0.847 | -7.62 | -0.00538 |
| New York | -0.0465 | -14.9 | 0.82 | -10 | 0.00648 |
| Alaska | -0.052 | -13.3 | 0.8 | -11.8 | -0.0133 |
| Connecticut | -0.0523 | -13.2 | 0.798 | -11.9 | -0.0584 |
| New Hampshire | -0.0549 | -12.6 | 0.789 | -12.7 | 0.0993 |
| American Samoa |  |  |  |  |  |
| Florida |  |  |  |  | 0.0411 |
| Guam |  |  |  |  | -0.0734 |
| Hawaii |  |  |  |  | -0.0135 |
| Kansas |  |  |  |  | 0.046 |
| Northern Mariana Islands |  |  |  |  |  |
| US Virgin Islands |  |  |  |  | -0.21 |

## Simulation of two second wave scenarios in Illinois

We simulate two scenarios of a second wave of the COVID-19 epidemic in Illinois using the model described in [2]. We calibrate our model using data through June 28 on hospital and ICU room occupancy by COVID-19 patients, the number of daily deaths of COVID-19 confirmed patients in hospitals, and the total number of daily deaths as publicly reported by the IDPH [6]. The results are shown in Figure 5. In the first scenario (shown in the top panel) we assume that the exponential growth during the second wave in Illinois would match the observed growth in other states that are currently experiencing a second wave. More specifically, we choose the relaxation factor *E* = 35% (to roughly match hospitalization growth rate in Texas). This choice is higher than the value *E*(TX) = 31% estimated using Eq. 2 to account for additional reduction in susceptible population fraction at the early phase of the second wave before hospitalizations start growing significantly. In this case, we confirmed that the exponential growth rate of hospitalizations (measured between August 1 and August 15) in our forecast is approximately 6.7%, matching the current observed growth in Texas.

**FIG. 5.**
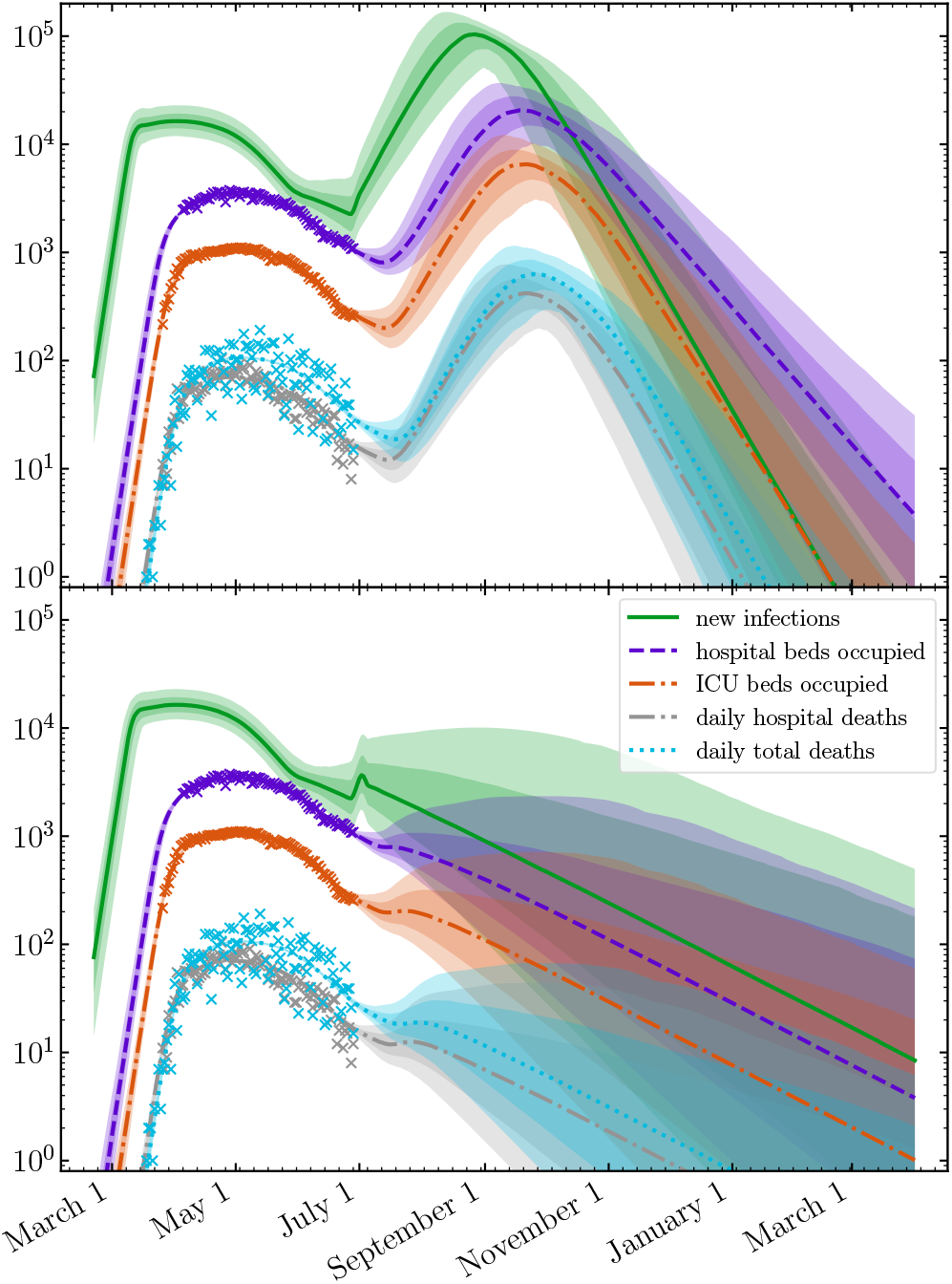
Top panel: the trajectory of the COVID-19 epidemic in Illinois following relaxation of mitigation starting July 1, leading to a second wave with the exponential growth rate as in Texas. This scenario corresponds to relaxation of mitigation by *E* = 35%. Bottom panel: the second wave dynamics in a scenario where the effective reproduction on July 5, 2020 (*E* = 0%). Different colors correspond to daily new infections (green), hospital occupancy (purple), ICU room occupancy (orange), hospital deaths (grey) and all deaths (cyan) predicted by our model [2] calibrated on existing data (crosses) up to June 28, 2020.

In the second scenario we assume mitigation steps carried out by the state of Illinois would be successful in reverting the effective reproduction number to the Phase 3 value of 0.92. We model this reversal as a transition taking place between July 5 and July 10. While we are unable to predict what changes in state policies would be sufficient to make this happen, we conjecture that setting strict limits to large gathering, and closing bars and indoor dining would bring superspreading under control. Indeed, a growing consensus has emerged that COVID-19, like previous coronavirus epidemics, is spread through large, rare events known as super-spreaders, wherein 80% of the transmission arises from only 10-20% of the infected cases [7–10]. These are driven by pre- or asymptomatic individuals [11], and throughout the world have been associated with airborne transmission especially in indoor environments [12] such as bars and restaurants [8, 13, 14]. These environments are crowded and are often marked by people talking loudly and removing their masks in close proximity, and the average length of stay at these events may be an hour or more. As a result, there is a high probability of infection. Some studies [12] have shown that closed indoor environments can be around 20 times more conducive to spread of COVID-19 than open-air settings. Anecdotally, bars, indoor dining, and even unofficial large parties are occurring within Illinois and other states [15] and can cause businesses to close through multiple secondary infections [16].

## Conclusions

The amplitude of the second wave in Illinois (triggered by the transition to Phase 4) that matches the exponential growth currently observed in Texas (relaxation of mitigation by *E* = 35% towards no-mitigation maximum) would significantly exceed the amplitude of the first wave. Both hospital and ICU room capacities statewide would likely be exceeded in the Fall of 2020. Conversely, under a scenario in which State-mandated mitigation of superspreader events reverts virus dynamics to Phase 3 (*E* = 0%), the second wave would be successfully prevented, and the hospital and ICU room capacities of the state would never be breached.

## Data Availability

Data is public

https://github.com/uiuc-covid19-modeling/pydemic

## ACKNOWLEDGMENTS

We gratefully acknowledge discussions with Mark Johnson at Carle Hospital, Sarah Cobey at University of Chicago, and Jaline Gerardin at Northwestern University. Our calculations would have been impossible without the data kindly provided by the Illinois Department of Public Health through a Data Use Agreement with Civis Analytics. This work was supported by the University of Illinois System Office, the Office of the Vice-Chancellor for Research and Innovation, the Grainger College of Engineering, and the Department of Physics at the University of Illinois at Urbana-Champaign. Z.J.W. is supported in part by the United States Department of Energy Computational Science Graduate Fellowship, provided under Award No. DE-FG02-97ER25308. This work made use of the Illinois Campus Cluster, a computing resource that is operated by the Illinois Campus Cluster Program (ICCP) in conjunction with the National Center for Supercomputing Applications (NCSA) and which is supported by funds from the University of Illinois at Urbana-Champaign. This research was partially done at, and used resources of the Center for Functional Nanomaterials, which is a U.S. DOE Office of Science Facility, at Brookhaven National Laboratory under Contract No. DE-SC0012704.

Our model is implemented in the open source Python 3 [17] package pydemic. The source code for pydemic is freely available online at https://github.com/uiuc-covid19-modeling/pydemic. This work made use of NumPy [18], SciPy [19], pandas [20], emcee [21], corner.py [22], and Matplotlib [23].

## Footnotes

1 The 5 phases of the Restore Illinois Project are described at [1]. Transition to Phase 4, which took place state-wide on June 26, 2020, is potentially especially dangerous since it involves opening up indoor seating in bars, restaurants, etc.—a major risk factor in spreading of COVID-19.

2 The Illinois Department of Public Health [3] includes positivity rate below 10% as one of several targets that counties should strive to achieve. Exceeding this target rate along with at least one other target would elevate the epidemic risk rate in a county.

